# Penalized longitudinal mixed models with latent group structure, with an application in neurodegenerative diseases

**DOI:** 10.1101/2020.11.10.20229302

**Authors:** Farhad Hatami, Konstantinos Perrakis, Johnathan Cooper-Knock, Sach Mukherjee, Frank Dondelinger

## Abstract

Large-scale longitudinal data are often heterogeneous, spanning latent subgroups such as disease subtypes. In this paper, we present an approach called *longitudinal joint cluster regression* (LJCR) for penalized mixed modelling in the latent group setting. LJCR captures latent group structure via a mixture model that includes both the multivariate distribution of the covariates and a regression model for the response. The longitudinal dynamics of each individual are modeled using a random effect intercept and slope model. Inference is done via a profile likelihood approach that can handle high-dimensional covariates via ridge penalization. LJCR is motivated by questions in neurodegenerative disease research, where latent subgroups may reflect heterogeneity with respect to disease presentation, progression and diverse subject-specific factors. We study the performance of LJCR in the context of two longitudinal datasets: a simulation study and a study of amyotrophic lateral sclerosis (ALS). LJCR allows prediction of progression as well as identification of subgroups and subgroup-specific model parameters.

## 1. Introduction

Longitudinal designs play a key role in biomedical research. In these studies, repeated measurements of the same quantities enable the study of temporal processes such as disease progression. Contemporary large-scale longitudinal datasets may include large numbers of observed variables, and are often heterogeneous, spanning multiple data subgroups such as disease subtypes. This leads to a subgroup structure that is often latent. In this situation, classical longitudinal models may be confounded by the latent group structure, or may simply be inapplicable due to the number of covariates.

In this paper, we propose an approach called *longitudinal joint cluster regression* (LJCR) for the heterogeneous data case that extends classical mixed modelling via a regularised mixture framework. In summary our approach posits models specific to latent subgroups indexed by *k*, i.e.:

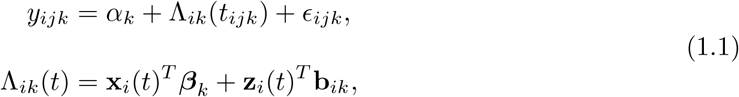

where *y*_*ijk*_ is the response for subject *i* at measurement *j* in subgroup *k, α*_*k*_ is the subgroup-specific intercept, *ϵ*_*ijk*_ are the (usually Gaussian-distributed) residuals. The term Λ_*ik*_(*t*) captures the time-dependent dynamics, with **x**_*i*_(*t*) the vector of covariates, ***β***_*k*_ the group-specific fixed effects, **z**_*i*_(*t*) the time-dependent covariates and **b**_*ik*_ the subject-specific random effects.

The subgroup-specific model parameters are a key feature of our model, which allows both the temporal dynamics, such as rate of progression, as well as the regression parameters ***β***_*k*_ to differ between subgroups. LJCR estimates *K* models of the form (1.1). The subgroup labels are treated as latent, which allows us to cope with the situation where the subgroup structure is entirely unknown at the outset.

LJCR is thus a joint modelling approach aimed at capturing heterogeneous longitudinal dynamics of disease progression by combining clustering, regression and linear mixed modelling. As described in detail below, LJCR considers both the distribution of **Y**|**X** and the distribution of **X**. This is done within a mixture framework, extending recent work by Perrakis et al. [2019] to the mixed model setting. Like Perrakis et al. [2019], we employ a joint cluster regression approach with regularization, allowing for group/cluster-specific regression parameters via a latent variable model. To deal with longitudinal dynamics, we incorporate a linear mixed effects intercept and slope model, and we develop a combination of L1 and L2 penalization and an efficient inference method to deal with small n and moderate-to-large p scenarios. Our method treats both the outcome variable and the explanatory variables as random quantities whose covariance matrix can be estimated. To find the optimal number of latent clusters within a given range, we employ a heuristic based on the elbow technique [Joshi and Nalwade, 2013].

Our work is motivated by challenges in longitudinal data analysis in the study of neurodegenerative diseases (NDDs). These diseases have complex underlying aetiology and display considerable heterogeneity in presentation and progression. Furthermore, as for many complex diseases in neurology and psychiatry, disease subtyping remains an open area of investigation. Hence one cannot typically assume that all subjects in a given study follow the same distribution, nor that subgroups are known at the outset. In general, NDD patients are characterized by heterogeneous progression profiles, leading to very different disease trajectories and increases in impairment that progress at different time-scales for each individual. This effect is particularly striking in motor neurone disease, or amyotrophic lateral sclerosis (ALS), a disease targeting the voluntary motor neurons. Most people with ALS succumb to the disease within 2-4 years, but around 10% of affected people survive more than 10 years [Swinnen and Robberecht, 2014]. The drivers behind these differences in progression remain incompletely understood. It is therefore imperative to develop computational methods that can infer underlying subgroup structure from observed data and leverage this to predict long- and short-term progression.

Modelling heterogeneous data is an active area for statistical research. For example, Dondelinger et al. [2020] used a joint penalized regression approach for estimation of high-dimensional fixed effects in heterogeneous data, but their approach is for cross-sectional rather than longitudinal data, and does not include random effects. Graphical models for heterogeneous data have also been considered in the literature [Danaher et al., 2011]. Various statistical models have been developed to infer both regression and group structures (latent variables); for example using regularized or unregularized mixture models [McLachlan], such as in [Khalili and Chen, 2007, Städler et al., 2010] where regularized mixtures of regressions were employed. Alternatively, [Xu et al., 2015] developed a multi-task approach using regularized LU-decomposition to map individual-specific models from *k* latent base models, where each individual-specific model is a linear combination of the base models. This approach, although flexible, is less convenient for identifying well-defined groups. Suresh et al. [2018] developed a deep learning multi-task model based on a LSTM (Long short-term memory) architecture. Patient groupings were first learned by clustering the embeddings of an autoencoder with LSTM structure using a standard Gaussian mixture model. Then a second neural net with a common LSTM layer and group-specific dense hidden layers was used to produce predictions for each patient. As this model is highly non-linear, interpreting the influence of specific variables becomes difficult.

An alternative approach is to extend conventional clustering approaches to the longitudinal setting. For instance, the k-means method for longitudinal data [Genolini and Falissard, 2010] is an implementation of k-means specifically designed to clustering longitudinal data. For a given number of clusters (*K*), the algorithm determines a clustering of individuals, where the progression scores **y**_*i*_ = (*y*_*i*_(*t*_*i*,1_), …, *y*_*i*_(*t*_*i,n*_))^*T*^ at time points *t*_*i*,1_ to *t*_*i,n*_ are treated as observations for each individual, and the Euclidean distance with Gower correction is used to optimise the cluster assignments. Note that this method does not take any covariates into account, and will not work very well in cases where none of the observation times coincide across individuals. Another consideration is how to detect the optimal number of clusters (subgroups or subtypes) using such methods [Everitt et al., 2001]. Various efforts have been made, either using nonparametric [Ray and Turi, 1999, Davies and Bouldin, 1979] or parametric approaches [Hurvich and Tsai, 1989, Schwarz et al., 1978].

None of the models described above take the distribution of the features **X** into account. Mixture regression models propose a mixture approach for the conditional distribution of **Y**|**X** and hence solely deal with the relation between the response variable **Y** and feature matrix **X**, disregarding any signal that would arise from the distribution of **X** itself. This would make it difficult to predict the response value (for example progression of a disease) on new intakes (patients) with new design matrix **X**^*∗*^, as we cannot assign these patients to a specific group, and would have to average across all possible groups. Motivated by this gap, Perrakis et al. [2019] extended the framework of mixture regressions to include the distribution of the features in the estimation of the latent group membership variable. Our work builds on this to incorporate longitudinal dynamics. Via a combination of the profile likelihood [Pinheiro and Bates, 2000] and some simple linear algebra, we show how an efficient Expectation-Maximisation algorithm can be developed, allowing for scaling to large *p* scenarios.

The remainder of the paper is organised as follows. We first present the methodological framework of our method, before describing the results of an in-depth simulation study to characterise its performance. We then apply our method to a real-world dataset of ALS patients, and analyse both the predictive performance of our model, as well as the properties of the inferred groups and the main features associated with longitudinal progression

## 2. Methodology

Our model consists of a joint mixture regression model similar to the one described in Perrakis et al. [2019], but where the fixed effects regression model is replaced with a mixed effects model with a random intercept and slope term, as first explored in Laird and Ware [1982], to represent the longitudinal dynamics. In order to deal with small subgroup sizes, we apply l2 regularisation to the fixed effects, which requires the development of an efficient inference algorithm based on the profile likelihood [Pinheiro and Bates, 2006]. For ease of exposition, we first describe the linear mixed model and inference in Subsection 2.1 for the case where the subgroups are known. We then describe the mixture model in Subsection 2.2 and present the full expectation-maximization (EM) algorithm for inference in the combined model in Subsection 2.3.

We have *M* observational units (usually patients or study subjects). Suppose **y**_*i*_ = (*y*_*i*_(*t*_*i*,1_), …, *y*_*i*_(*t*_*i,n*_))^*T*^ denotes an *n*_*i*_-dimensional vector of responses at time points **t**_*i*_, and **X**_*i*_ denotes an *n*_*i*_ × *p* matrix of observed covariates for observational unit *i* ∈ {1, …, *M*}. Let *k* ∈ {1, …, *K*} denote the group label and *z*_*i*_ ∈ {1, …, *K*} represent the true (latent) group label indicator for the sample (**y**_*i*_, **X**_*i*_) with *p*(*z*_*i*_ = *k*) = *τ*_*k*_. Let **y** be the stacked response vector of length 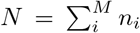 collecting the **y**_*i*_, and let **X** be the stacked design matrix collecting the **X**_*i*_. Define 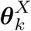 and 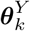 to be the group-specific parameters; respectively parameterizing the marginal distribution of **X** and the regression model of **y** on **X**.

### 2.1 Linear mixed effects model for longitudinal dynamics

We model the longitudinal dynamics of the outcome variable *y*_*i*_ with a mixed effects model:

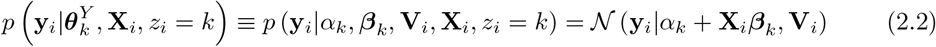

In other words, we introduce a dependency between longitudinal observations of patient *i* via the covariance matrix **V**_*i*_. If **V**_*i*_ is diagonal with diagonal elements 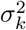 then a fixed effects model is recovered.

We define *V*_*i*_ using a standard longitudinal mixed model approach with random effects for the intercept and slope (Laird and Ware [1982]). Conceptually, we model 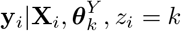 as

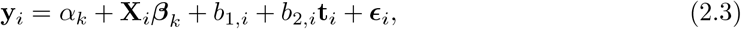

where **b**_*i*_ = (*b*_1,*i*_, *b*_2,*i*_)^*T*^ *∼ 𝒩* (0, **D**_*k*_) and 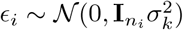. Here 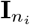 is the *n*_*i*_ × *n*_*i*_ identity matrix. Note that **b**_*i*_ implicitly depends on *z*_*i*_ = *k*; we chose not to make this explicit in the notation to avoid a redundant subscript. It follows that

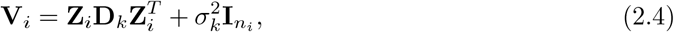

where **Z**_*i*_ = (1, **t**_*i*_) is the *n*_*i*_ × 2 design matrix of random effect covariates (in our case, an intercept and the observation time variable). The notation introduced in eqs. (2.2-2.4) differs slightly from the more standard notation for longitudinal models in eq. (1.1), but will simplify exposition in what follows.

Our method needs to be robust to low sample sizes and large numbers of covariates. We use *L*_2_ penalization to regularize the model and allow for efficient estimation of the fixed effect parameters. In the following, we drop the latent group indicator *z*_*i*_ = *k* and assume that the group labels are known.

The linear model for the response **y**_*i*_ can then be written as:

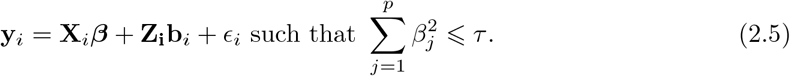

In general, our model will include an intercept term, as in eq. (2.3); to simplify notation we assume that this has been integrated into the design matrix as an additional column of ones. The corresponding objective function takes the form:

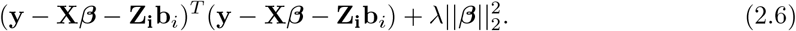

Straightforward optimization of this objective function would involve inverting *M p* × *p* matrices (where *M* is the number of patients). For large *M* and *p* this is not computationally feasible. Instead, we describe a more computationally efficient approach using the QR decomposition.

We follow Pinheiro and Bates [2000] in augmenting the vectors **y**_*i*_ and design matrices **X**_*i*_ and **Z**_*i*_ using a pseudo-data approach. First, note that the likelihood has the form

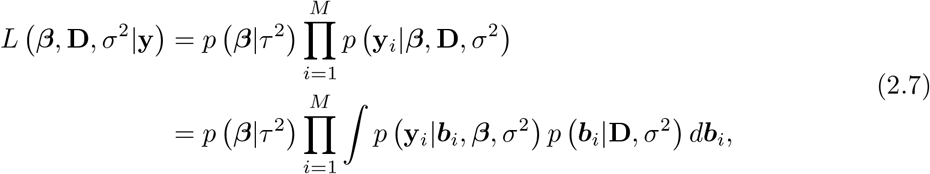

where the *p*(***β***|*τ*^2^) term is a multivariate normal prior with variance *τ* ^2^ inducing a ridge penalization with parameter *λ* = *σ*^2^*/τ* ^2^. Let us parameterise the multivariate normal distribution for **b**_*i*_ as

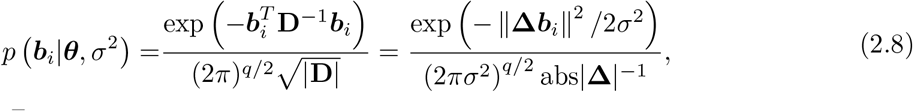

where *σ*^2^**D**^−1^ = **Δ**^*T*^ **Δ**, and ***θ*** denotes the free parameters in **D** (or equivalently **Δ**). Note that in our case the number of random effect parameters *q* = 2. We can define augmented vectors and design matrices,

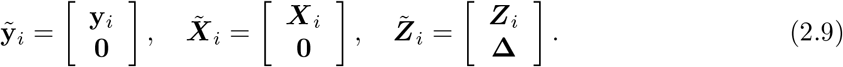

Pinheiro and Bates [2000] show that given an estimate of 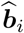, we can express the likelihood (without the penalizing prior on ***β***) as

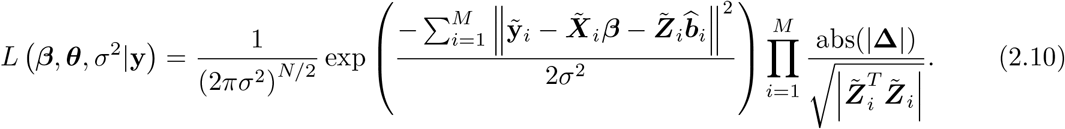

Now, we can show that with the prior on ***β***, we get:

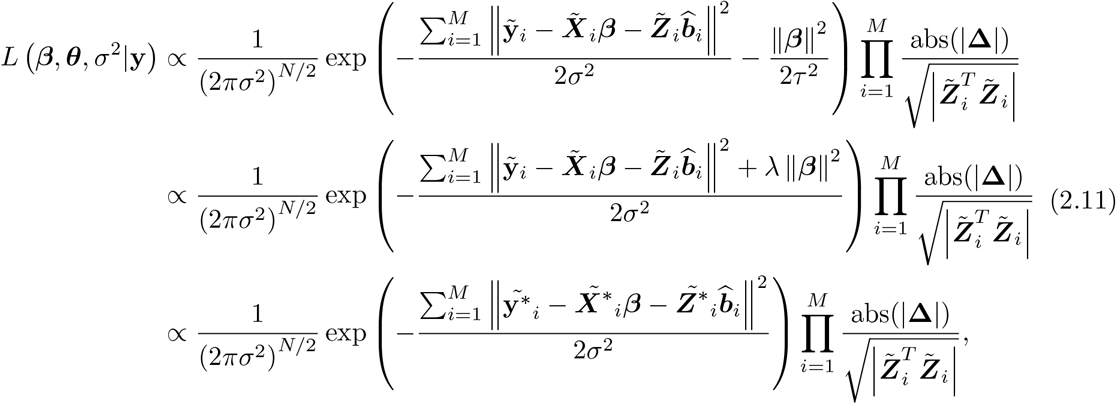

where we have further augmented the vectors and matrices as

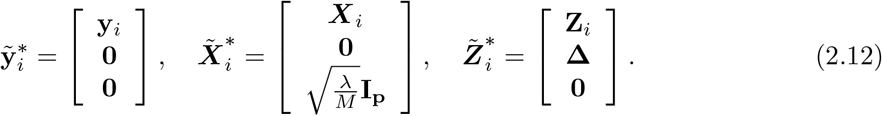

Following Pinheiro and Bates [2000], we can work out that the profiled likelihood is

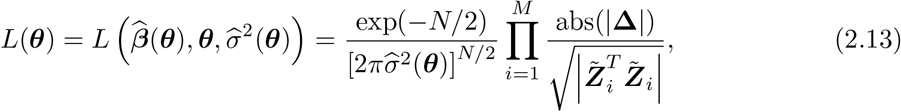

with 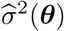 defined by the residual sum-of-squares. Instead of the naive approach of first estimating 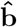 and 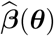 in order to get 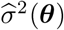, we can more efficiently calculate the latter via the QR decomposition:

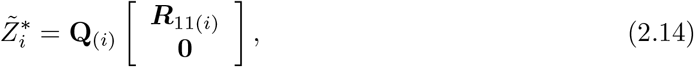

where **Q**_(*i*)_ is (*n*_*i*_ + *q* + *p*) × (*n*_*i*_ + *q* + *p*) and ***R***_11(*i*)_ is *q* × *q*. In our case, *q* = 2, therefore

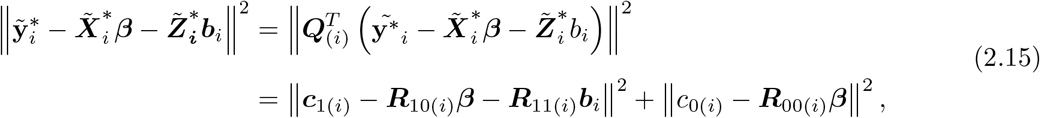

where

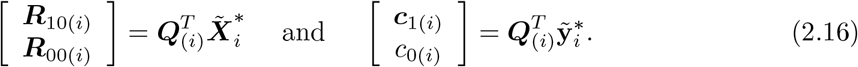

By integrating out the **b**_*i*_, eq. (2.11) can then be shown to become

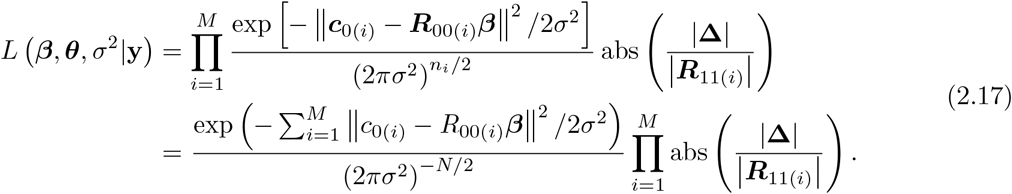

The term in the exponent is a residual sum-of-squares across patients *i*, which can be calculated using an additional QR decomposition as follows:

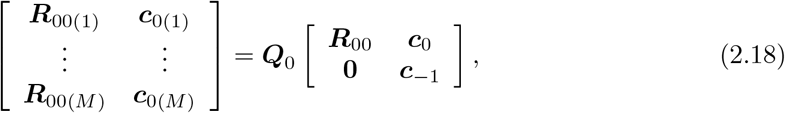

which leads to:

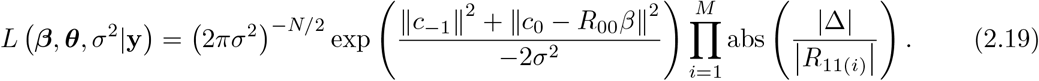

Using the maximum likelihood estimates for ***β*** and *σ*^2^:

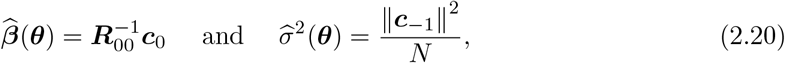

we get the final expression for the profile likelihood:

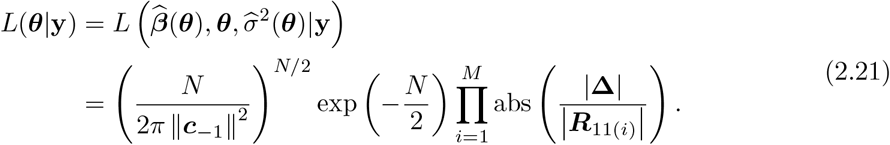

We are now able to optimize eq. (2.21) with respect to ***θ*** = *D* and then use the maximum likelihood estimate 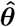 in eq. (2.20) to get the estimates for 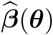 and 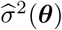. Note that this approach only involves a single matrix inversion of ***R***_00_; however, since this is an upper triangular matrix, we can simply solve for 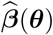 by forward substitution in the equation 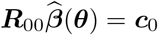.

Finally, we note that the QR decomposition in eq. (2.18) is rather inefficient for the high-dimensional case, because of the inflation of the starred matrices with *p* penalty terms, which leads to having to calculate the QR decomposition of a matrix of size (*M* ∗ (*n*_*i*_ + *p*)) × (*p* + 1). We can avoid the computational burden by first noting that the matrix on the left-hand side in eq. (2.18) is taller than it is wide. Let us define:

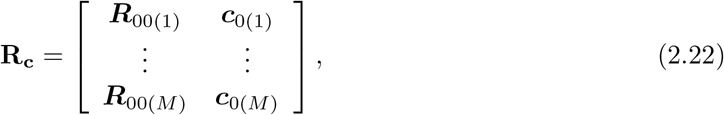

and

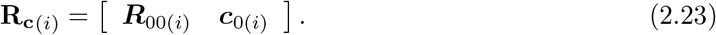

Then it can be shown that the R-matrix from the QR decomposition of **R**_**c**_ can be obtained by Cholesky decomposition of the cross-product **A** = **R**_**c**_^*T*^ **R**_**c**_. But the cross-product of the (*M* ∗ (*n*_*i*_ + *p*)) × (*p* + 1) matrix **R**_**c**_ is just the sum of *M* cross-products 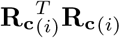. We can further optimize the calculation by noting that each 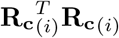 is of the form:

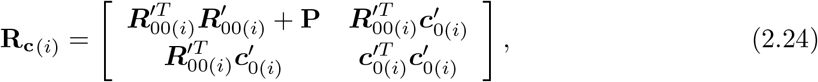

where 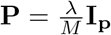, and 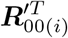 and 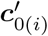 result from transforming the unpenalized vector and matrix 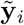 and 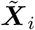defined in eq (2.9) by the upper left (*n*_*i*_ + *q*) × (*n*_*i*_ + *q*) matrix of 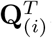, similarly to eq. (2.16). As a result, we can avoid calculating the cross-products of (*n*_*i*_ + *p*) × (*p* + 1) matrices in favour of *n*_*i*_ × (*p* + 1) matrices, followed by adding **P** to the upper left *p* × *p* matrix.

### 2.2 Mixture model for latent group structure detection

We are now ready to define the mixture model that combines a latent group membership variable with the longitudinal regression model defined in Subsection 2.1. Conditional on *z*_*i*_ = *k*, i.e. knowing the cluster memberships, the joint likelihood of (**y**_*i*_, **X**_*i*_) can be decomposed into the mixed effects regression model for **y**_*i*_|**X**_*i*_, and a multivariate model for **X**_*i*_ as follows:

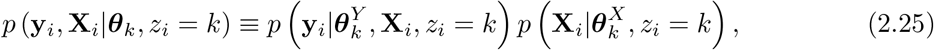

where 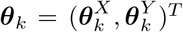. Marginalizing out the latent variables leads to a mixture regression of the form:

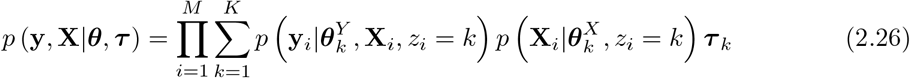

where ***θ*** = (***θ***_1_, …, ***θ***_*K*_)^*T*^ and ***τ*** = (***τ*** _1_, …, ***τ*** _*K*_)^*T*^. The model formulation presented in eq. 2.26 has been studied by Ingrassia et al. [2012] under the non-longitudinal setting and in the context of ML estimation.

*Gaussian graphical model for* 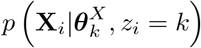. Throughout this paper we assume that covariates can be modelled via *p*-dimensional multivariate Gaussian distributions such that 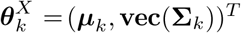, where ***µ***_*k*_ is the mean and **Σ**_*k*_ is the *p* × *p* covariance matrix. To mitigate computational costs during inference, we do not attempt to model the time-dependencies for time-varying covariates, but instead asssume that the overall mean and covariance are sufficiently representative of the underlying phenotype that we want to capture via the mixture components. To deal with the potentially large number of parameters in **Σ**_*k*_, we use regularization via the graphical lasso introduced in Friedman et al. [2008] (package glasso in **R**, Friedman et al. [2015]). The graphical lasso induces sparsity in the inverse covariance matrix, denoted by 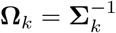 for group *k*, where we set the graphical lasso penalty to be *–ξ*‖**Ω**_*k*_‖_1_, in such a way that *ξ >* 0 controls the strength of regularization and ‖.‖_1_ is the *L*_1_ norm. Then for known group labels the graphical lasso estimate is given by solving the following maximization problem

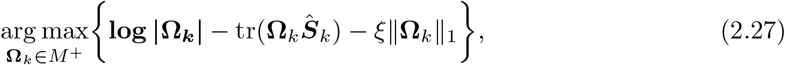

where *M* ^+^ is the space of positive definite matrices and ***Ŝ***_*k*_ is the ML covariance estimate of *X*_*k*_. In practice, this will be weighted by the responsibilities *m*_*ik*_ = *p*(*z*_*i*_ = *k*|**y**_*i*_, **X**_*i*_, ***θ***_*k*_) of each patient, using ***ℳ***_*k*_, the *N* × *N* diagonal matrix with entries ***ℳ***_*k*_(*r, r*) = *m*_*ik*_ if row *x*_*r*_ corresponds to covariates for patient *i*. The empirical estimate for the covariance matrix becomes ***Ŝ***_*k*_ = ***X***^*T*^ ***ℳ***_*k*_***X***. If the covariates are not time-varying, then for the purpose of eq. (2.27) the matrix ***X*** can be simplified to an *M* × *p* matrix without loss of generality.

*Linear mixed model for* 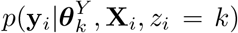. The regression term 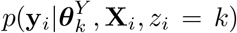 corresponds to the mixed effects model in eq. (2.2). However, in the case where the latent variable *z*_*i*_ is unobserved, we need to additionally account for the responsibilities *m*_*ik*_ of each patient. In other words, the likelihood function for group *k* is of the form

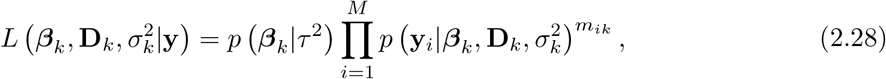

which means that eq. (2.17) can be shown to become

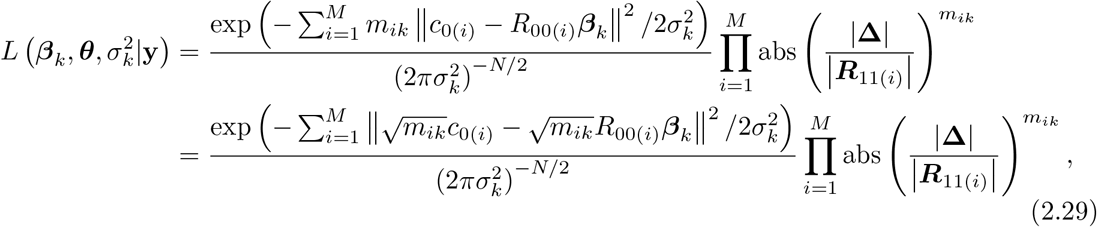

and consequently the matrix that needs to undergo QR decomposition becomes:

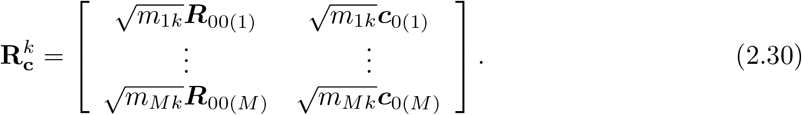

Note that 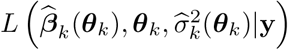 can be optimized individually for each *k*, leading to potential efficiency gains using parallel computation.

### 2.3 Expectation-Maximization Algorithm

Inference of ***β***_*k*_, **D**_*k*_ and *σ*_*k*_ for each group *k* ∈ *K* is complicated by the fact that the group indicator variables *z*_*i*_ are unobserved. We employ an expectation-maximization (EM) algorithm, similar to Perrakis et al. [2019], to perform this inference. We describe the initialisation, expectation (E-step) and maximisation (M-step) below; the algorithm is also summarized in Algorithm 1.

#### Initialisation

We initialise ***β***_*k*_ values across all groups, by running a simple penalised linear model using the `glmnet` package in R (Hastie [2020]). We initialise **D**_*k*_ and *σ*_*k*_ by choosing randomly generated positive definite matrices and scalar values, respectively, across all groups.

#### E-step

We estimate the responsibilities following Perrakis et al. [2019], using the the following formula:

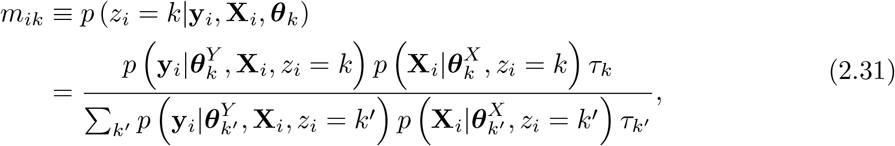

with the modification that in our work, *i* refers to patients rather than data points, and 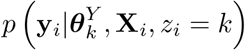 is defined as in eq. (2.2).

In the next step, which is now the M-step, we need to optimise the objective function stated

in eq. (2.28) (which comes in the form a profile log-likelihood function) and update 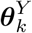 and 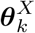 using the responsibilities *m*_*ik*_ from the E-step. For 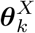, let **W**_*k*_ = ***ℳ***_*k*_**X** be the weighted covariate matrix, then 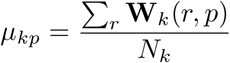 is the update for *µ*_*k*_, where *N*_*k*_ = ∑_*i*_*m*_*ik*_. **Σ**_*k*_ can be updated using the graphical lasso update in eq. (2.27). For 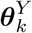, we update ***β***_*k*_, **D**_*k*_ and *σ*_*k*_ using the profile likelihood in eq. (2.21).

Algorithm 1 shows the pseudo-code of the EM procedure deployed in the LJCR algorithm. Here *objective function* refers to the eq. (2.28) and ***ℳ***_*k*_ is the *N* × *N* matrix containing the responsibilities on the diagonal and zero elsewhere. The maximum number of iterations *N*_*it*_ is set to 100, although the loop may terminate early if we reach convergence, or if the size of one of the groups (*N*_*k*_) gets very small. This latter condition is necessary to avoid splitting individuals into very small groups where estimation of ***β***_*k*_, **D**_*k*_ and *σ*_*k*_ would not be reliable.

##### Algorithm 1: Pseudo-code of the utilized EM algorithm

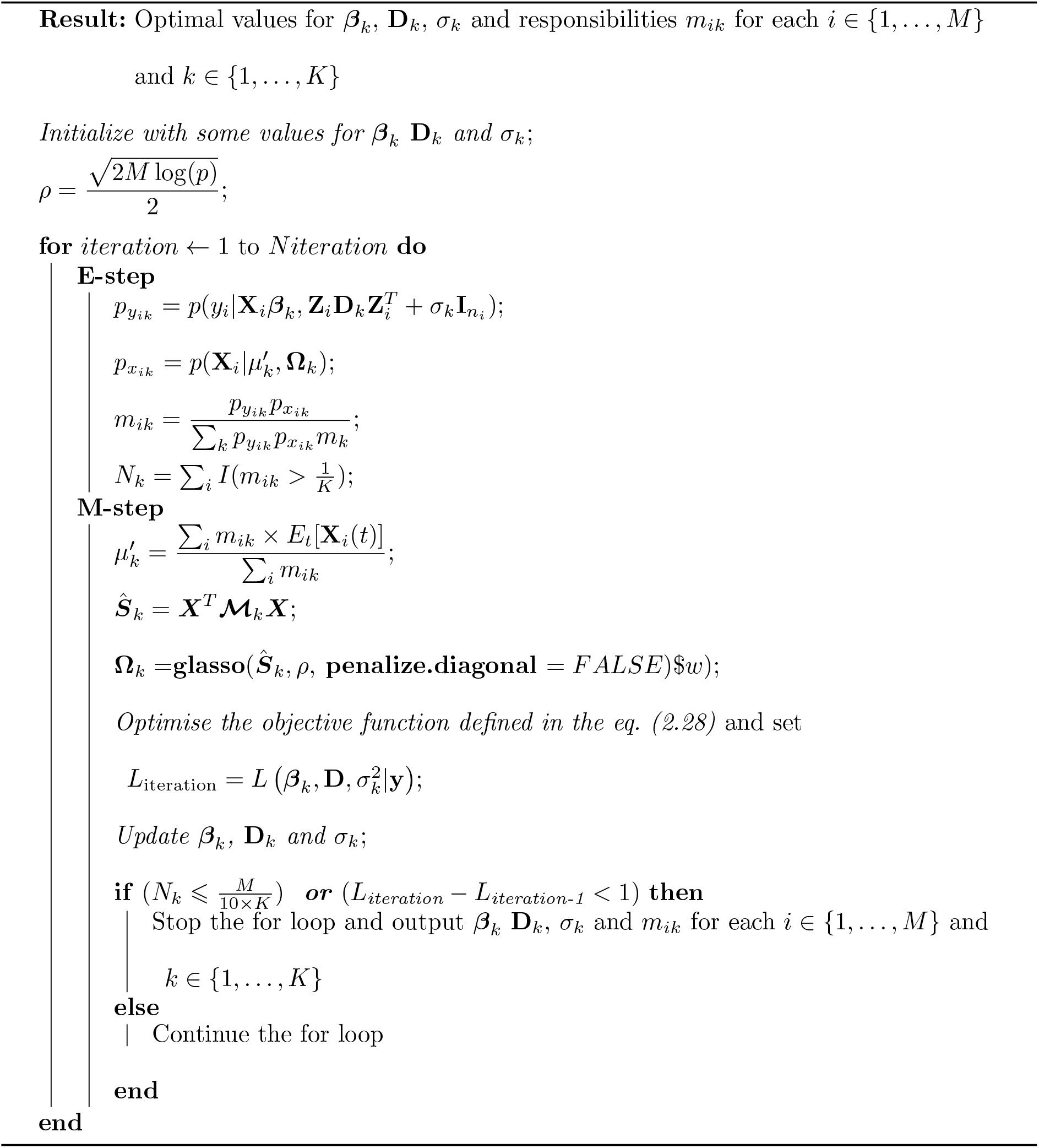

## 3. Results

We apply the LJCR model to two different longitudinal datasets; a simulated dataset, which allows us to evaluate the ability of the model to recover known subgroups and regression coefficients; and the PRO-ACT dataset (Atassi et al. [2014]), the largest database of clinical trials of patients with ALS (Amyotrophic Lateral Sclerosis). The latter allows us to both evaluate the predictive performance of the model on a real-world dataset, and to gain novel insights into factors that may be associated with different clinical phenotypes and progression trajectories in this disease.

### 3.1 Simulation Study

We perform a simulation study to test the performance of the LJCR method under a range of scenarios. More precisely, we study how well the model can predict the three parameters ***β***_*k*_ (coefficients for the fixed effects), *σ*_*k*_ (variance), and **D**_*k*_ (covariance matrix of the random effects) under scenarios with different number of individuals *M* and number of covariates (features) *p*. We then compare the performance of the LJCR algorithm with three different methods as follows:

- kml: k-means method for longitudinal data [Genolini and Falissard, 2010] (package kml in R).
- Clustering+LMM: In this method we first use the standard k-means algorithm to cluster the data (based on the response values *y*) and then apply a Linear Mixed Effects Model (LMM) (Schafer [1998]) (package LMM in R by Zhao [2020]) on each cluster.
- Baseline method: Here we cluster the data as above and then apply a simple linear model with a random slope only, without including the fixed effects for the design matrix **X**. This provides a convenient baseline that is not affected by the dimensionality of **X**.

Here we generate 10 sets of independent datasets where in each of those sets we generate 4 different datasets (in total 40) with the dimension of (*M* × *n*_*i*_) × *p* where *M* = 400 (number of individuals), *n*_*i*_ = 10 (number of observations/data points for each individual *i*), and *p* ∈ {100, 500, 1000, 7000} (number of features/covariates). We use these set of generated data to show the performance of the LJCR model when varying the number of features *p* (see figure 1). We then generate another 10 sets of independent datasets where in each of those sets we generate 4 different datasets (in total 40) with the dimension of (*M* × *n*_*i*_) × *p* where *M* ∈ {500, 1000, 4000}, *n*_*i*_ = 10, and *p* = 7000. Again, we use these set of generated data to show the performance of the LJCR model when varying the sample size *M* (see figure 1). The number of groups across all these simulation scenarios is assumed to be *K* = 3.

**Fig. 1:**
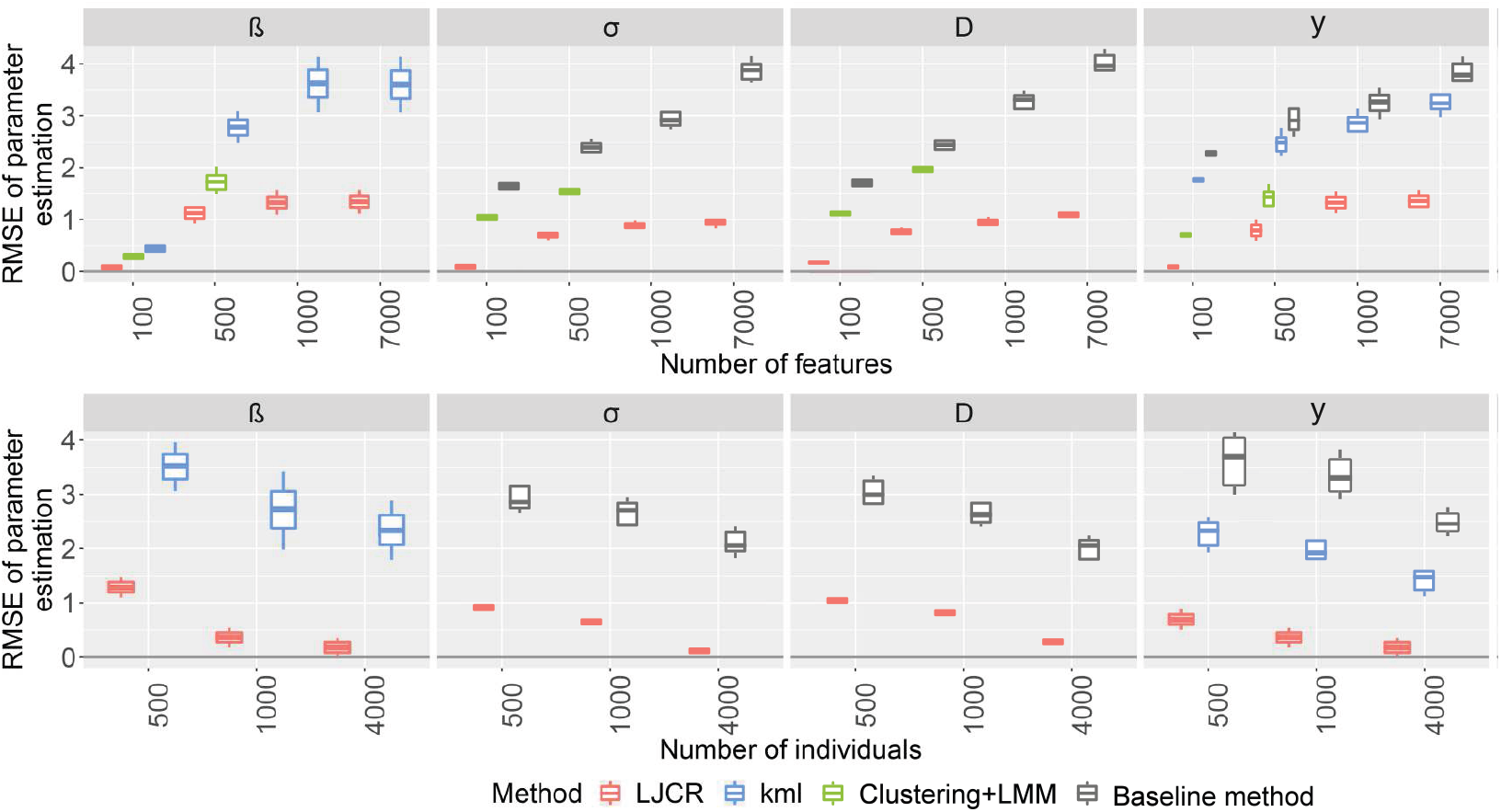
Comparison of the performance of the LJCR, k-means for longitudinal data, clustering and then running LMM on each cluster (Clustering+LMM), and baseline methods on simulated data when varying the number of features (top row), and sample size (bottom row). The boxplots represent the estimates of RMSE obtained from 10 independent simulated datasets. Missing boxplots for Clustering+LMM indicate situations where the dimensionality was too high to apply this method. Also note that the baseline method will not return an estimate for the ***β***_*k*_, and kml will not return an estimate for *σ*_*k*_ or ***D***_*k*_.

In both of the aforementioned sets of simulated data, we use normal distribution (using rnorm in R) with mean 0 and variance 1 to generate the data. We then sample the initial values for ***β***_*k*_, **D**_*k*_ and *σ*_*k*_ from a normal distribution with mean 0 and variance 1, randomly generated positive definite matrices (using genPositiveDefMat() in clusterGeneration R package by Weiliang Qiu [2015]), and uniformly generated scalar values, respectively, across all groups.

Figure 1 shows a comparison between the LJCR, the k-means for longitudinal data (Genolini and Falissard [2010]), clustering and then running LMM on each cluster (Clustering+LMM), and baseline methods. Performance of the all models have been tested on the generated datasets we explained before, varying the number of covariates and samples. Notice that in the kml method, we compute the mean trajectories of each group *k*, using the function calculTrajMeanC in the kml package, and consider them as the values of ***β***_*k*_. We then directly use the obtained ***β***_*k*_ values to calculate responses *y*_*i*_. We see that in the both scenarios where we increase the number of features *p* or the sample size *M*, the LJCR outperforms all the models in terms of prediction of both parameters ***β***_*k*_ and response values *y*_*i*_. We could not apply the Clustering+LMM method beyond *p* = 500, as multivariate nature and high-dimensional setting makes it infeasible. There is no covariate matrix **X** used in the baseline method, hence there would be no ***β*** obtained in this method.

Next we perform a sensitivity analysis of the LJCR model to look at the effect of different values of hyperparameters *λ* in eq. (2.6). We define a set of hyperparameters *λ* ∈ {0.1, 1, 10, 100} and allow the LJCR model to pick up the best value via 10-fold cross-validation. Figure 2a shows which hyperparameter values are chosen for different degrees of sparsity of the initial vector ***β***. Here we have *p* = 7000, *M* = 400 and *n*_*i*_ = 10 for all *i*. The test is performed for 10 simulated datasets. The size of each circle represents the number of times each value of *λ* is selected in a simulation run. As expected, when having highly sparse ***β***, then a large hyperparameter is chosen by the model. Figure 2b shows the relative ROC curve for different values of *λ* under the highly sparse scenario where 6500 out of 7000 values are near zero (93% sparsity). While high values of the hyperparameter naturally result in larger areas under the ROC curve (AUC) values, we note that even for misspecified *λ* values, the drop in AUC is modest.

**Fig. 2:**
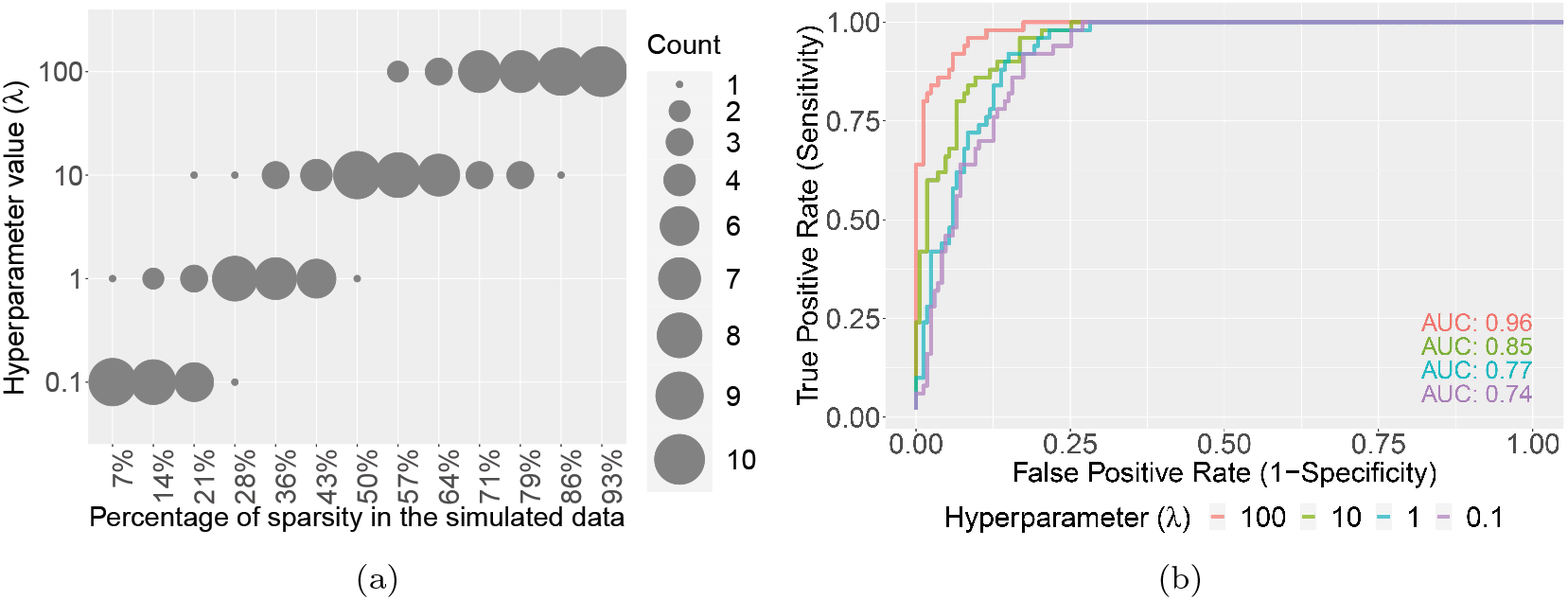
(a) The number of times the hyperparameter (*λ*) is selected by a 10-fold cross validation. The *x*-axis represents the proportion of sparse betas (everything below the 0.01 threshold) in the initial vector ***β*** that is used to simulate the data. (b) ROC curves of the estimated 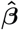 and associated AUC values for different hyperparameter values.

### 3.2 ALS PRO-ACT Data

PRO-ACT (Pooled Resource Open-Access ALS Clinical Trials) is a publicly available database containing industry and academic clinical trials of patients with ALS disease (Amyotrophic Lateral Sclerosis) (Atassi et al. [2014]). It is a longitudinal dataset including records of each individual across repeated visits to clinic. PRO-ACT is the largest ALS clinical trials database ever created, with more than 8500 patient records, including demographic and laboratory data, medical histories and functional scores.

In our study, we have used a subset of PRO-ACT collected longitudinally at different observation time-points. The response variable (*y*_*i*_) is the ALSFRS (ALS Function Rating Scale) score. This score captures the overall state of the disease and can be considered as a progression score for people living with ALS. The ALSFRS scale is a list of 10 different assessments of motor function (such as the ability to move an object, the ability to eat with cutlery, the ability to handwrite, etc.), with each measure ranging from 0 to 4, with 4 being the highest (normal function) and 0 being no function. The score for the individual questions are then summed together to generate the total ALSFRS score, which ranges between 0 − 40.

First of all, we have discarded all patients with single observation (one visit to the clinic). Then to deal with the missing values we interpolate using inter-subject sectional linear interpolation; i.e. we look at each individual *i* and replace every missing value with the average value between its previous and next observations/datapoints. We are then left with a cohort of *M* = 4821 patients and *p* = 55 observed features (covariates).

Supplementary table S1 shows all the *p* = 55 features used in the PRO-ACT dataset (Atassi et al. [2014]).

We have applied the LJCR algorithm to find the underlying latent subtypes.

Figure 3 shows that using the elbow technique (Joshi and Nalwade [2013]), *K* = 9 latent subtypes (groups or clusters) represents the optimal number of groups when applying the LJCR to the PRO-ACT dataset. The idea behind the elbow technique is to choose a number of clusters so that adding another cluster does not result in better model to fit to the data. More precisely, we look at the relative change of the values in each pair of consecutive clusters (gradient slope) and then compare the differences.

**Fig. 3:**
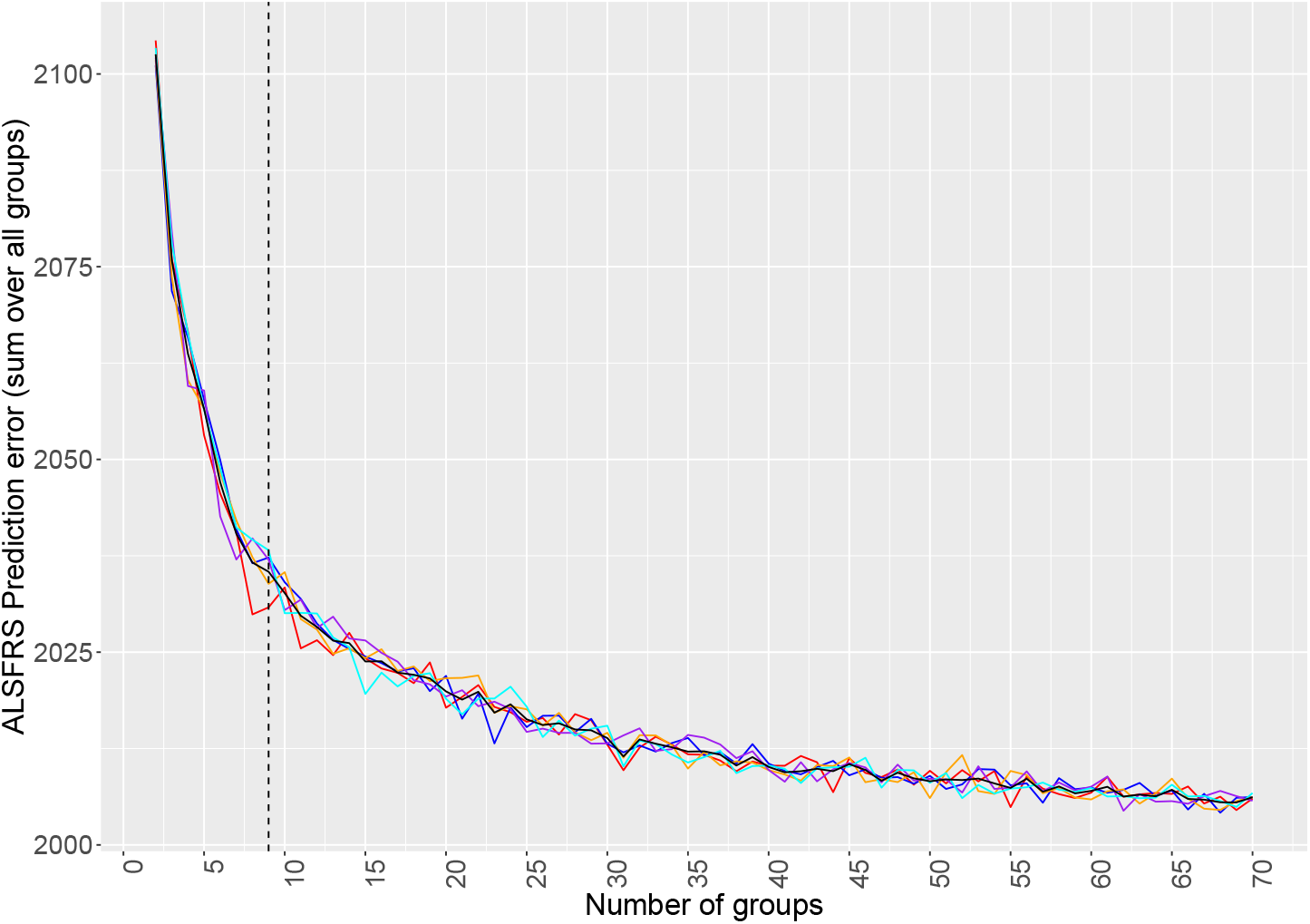
Performance of the LJCR model on the PRO-ACT ALS data when varying the number of latent subtypes (groups). The LJCR model is executed five times (shown in different colors) each time with different initial values for the parameters ***β***_*k*_, *σ*_*k*_, and **D**_*k*_. The number of latent groups varies with *K* ∈ {2, …, 70}). For each *K* (x-axis), the y-axis represents the mean prediction error for the ALSFRS scores across the groups. We calculate the error between the predicted response values (ALSFRS) (after running the LJCR and assigning the group membership) and the true ALSFRS values on the whole ALS PRO-ACT samples (training set). The black curve represents the mean of the colored curves. The elbow method (Joshi and Nalwade [2013]) is then used to identify *K* = 9 as the optimal number of subtypes for this dataset.

Figure 4a shows that there are 4 groups labels which contain most of the population size (*k* = {1, 5, 7, 9}). Figure 4b demonstrates that in the pre-mentioned group labels men are susceptible to be diagnosed with ALS disease earlier in age than women (about 2 − 5 years).

**Fig. 4:**
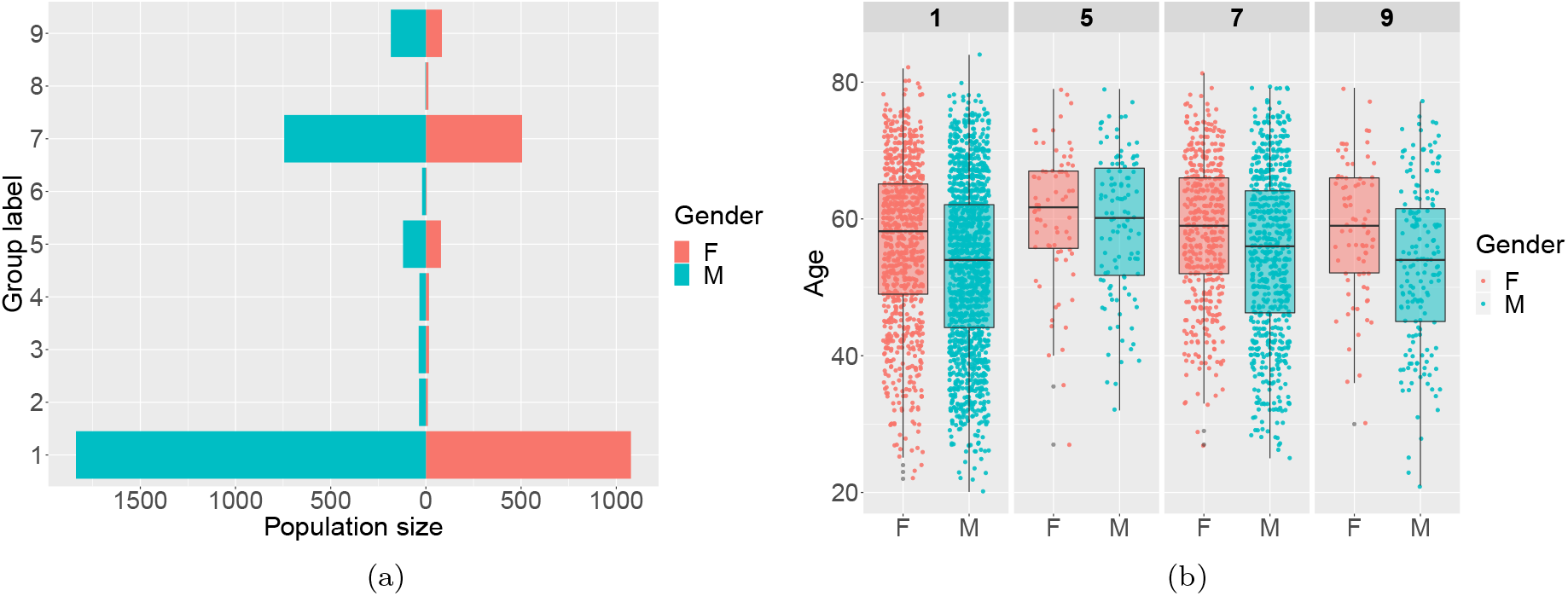
Application of the LJCR model to the PRO-ACT ALS study. (a) Pyramid plot showing the mixture component sizes. (b) Box-plots showing the age at baseline distribution for the groups with the largest population size *k* = {1, 5, 7, 9}, with the blue color standing for male and red for female.

Figure 5 shows the estimated effect size (***β***_*k*_) for each feature in group *k* ∈ {1, 5, 7, 9}. As an interesting result, we observe that the effect of Mean Corpuscular Hemoglobin Concentration (MCHC) has a positive effect size in group label 7, unlike in the other groups. Similarly, Absolute Basophil Count has a negative effect size in group label 7 which is in contrary to all other group labels.

**Fig. 5:**
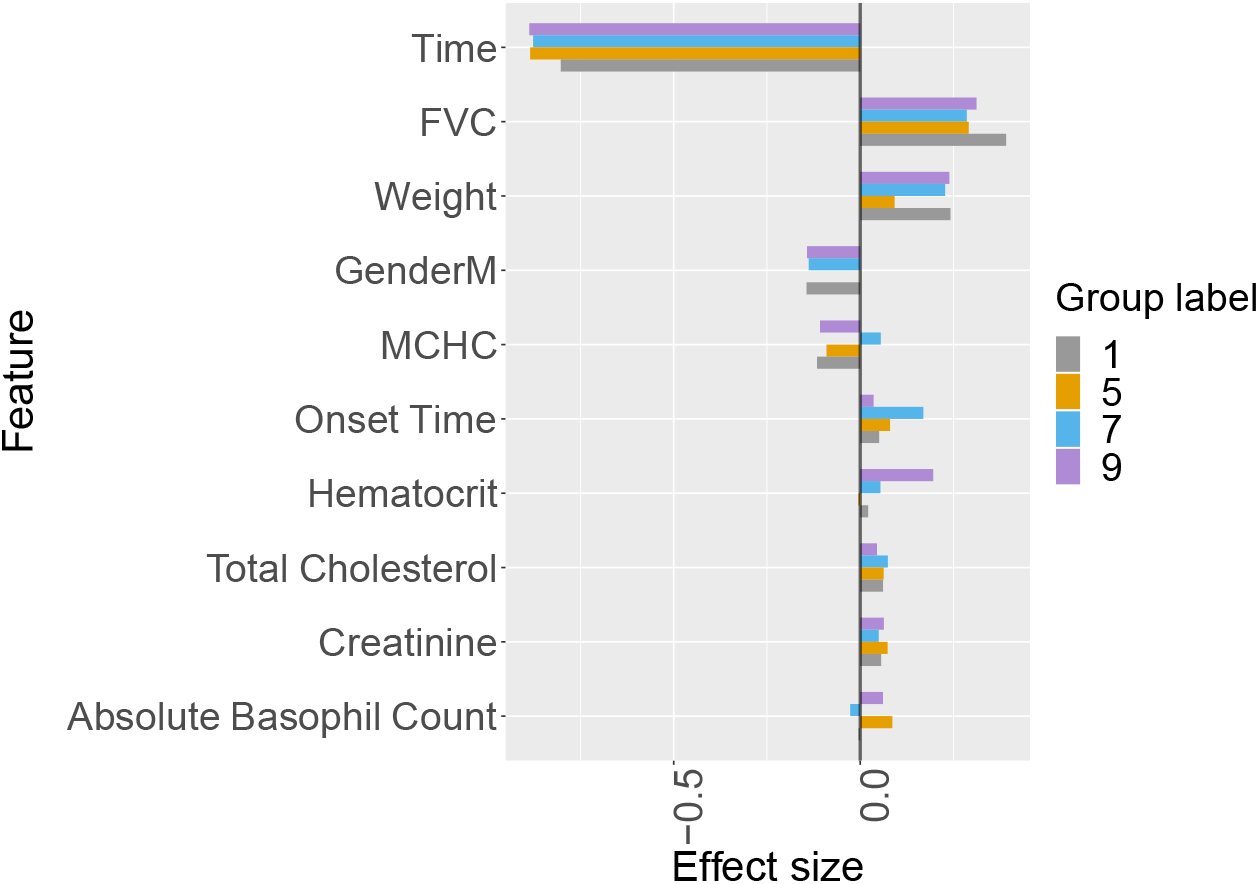
Application of the LJCR model to the PRO-ACT ALS dataset. The bar plots show the estimated effect size (estimated ***β***_*k*_ parameters) for the group labels with largest population size (*k* = {1, 5, 7, 9}). Note that MCHC stands for Mean Corpuscular Hemoglobin Concentration. Time refers to time on the study (in days, but scaled here to make the effect sizes comparable).

Many of these factors have been previously associated with the rate of ALSFRS decline including weight, FVC and age (Mandrioli et al. [2015]). Plasma creatinine has been previously associated with outcome in ALS and may act as a marker of muscle reserve (Mitsumoto et al. [2020]). The observation of different relative effect sizes between groups for cholesterol and weight suggests that cholesterol is not acting only as a proxy for weight. Indeed there is evidence that serum cholesterol may be an important marker of the extent to which a dysfunctioning motor system is energy deficient (Dupuis et al. [2008]). The observation that serum cholesterol may have a different relative effect in different patient groups is important because clearly hyperlipidaemia can be harmful in certain contexts and lead to, for example, cardiovascular disease; therefore it would be important to recommend dietary changes to boost cholesterol only when clinically appropriate. The observation that MCHC and hematocrit are associated with outcome is a novel finding. This could be interpreted in the context of resistance to respiratory failure however this does not explain the different direction of effect in groups 9 and 1. Equally the finding that absolute basophil count is a predictor of outcome is novel although peripheral immune cells have been linked to CNS inflammation and disease progression (Butovsky et al. [2012]). Discovering the differences in CNS inflammation between groups 9 and 5, where basophils have a positive correlation, and groups 1 and 7 where there is no correlation or a negative correlation, could guide personalised immunotherapy for ALS.

One potential issue with prediction of progression for new patients is that we only have access to the covariates **X**_*i*_ for assigning the new subject to an existing mixture component. In order to test whether this is sufficient, we perform an experiment where we first train the model on the whole dataset (4821 individuals) to assign each individual a group label. Then we choose a random set of 50 people as our test set and re-run the model on the remaining training set (4771 people). Finally, for each individual in the test, we find the estimated assigned group label by solving this problem: Find the group label with the highest probability of test individual *i* falling into that group based on the mean and variance of distribution of the observed feature training set (**x**_*ik*_) for each group label *k*. Here we applied the graphical lasso [Friedman et al., 2008] to calculate mean and covariance matrix components for each subgroups (**X**_*k*_). We have repeat this procedure 10 times. Figure 6 shows that the LJCR models performs reasonably well in detecting the same group labels that would have been assigned when training on both the response and the covariates.

**Fig. 6:**
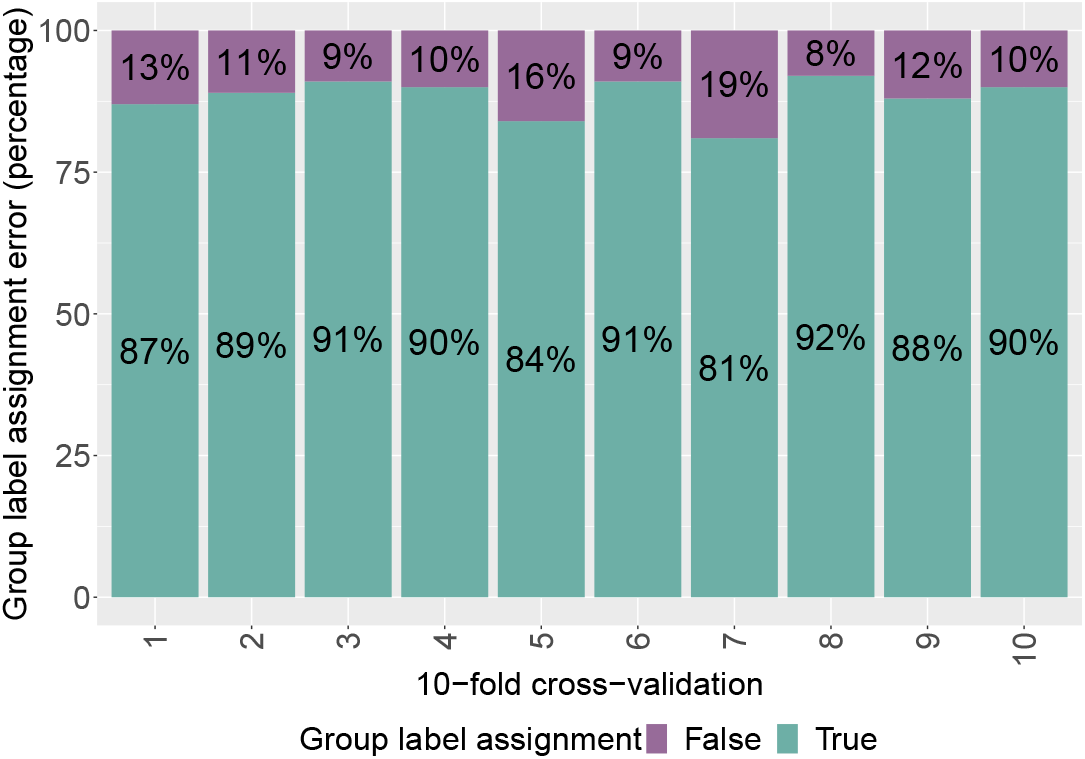
Performance of the LJCR model in detecting how well patients could be classified based on their covariates alone (no ALSFRS), compared to the group label that they were assigned when including both covariates and responses (ALSFRS) in the training of the mixture model. We train the model 10 times with different randomly selected test sets containing 50 individuals. The bar plots represent the percentage of group assignment error for each fold.

Figure 7a shows the density plots for the ALSFRS prediction error of the different mixture components. Note that group 1 is the largest, hence the MSE prediction error for the ALSFRS total score should be lower, since more data is available to estimate the parameters. Figures 7b shows a density plot where we have used the group labels inferred from the LJCR model, but then refit for each group using the LME model (linear mixed effects model). We see that this re-fitting after inference of the group memberships by the LJCR algorithm improves the prediction error.

Figure 8 shows the performance of the LJCR model in ALSFRS prediction under two scenarios; in the first scenario we test prediction for unseen individuals (Figure 8a) where we predict ALSFRS value of a test set of 50 new individuals. In the second scenario we test prediction for unseen time-points on individuals where the previous time points were included in the training set (Figure 8b). Here we predict ALSFRS scores at two time-points for 5 randomly selected individuals. Both figures 8a and 8b show that the prediction performance using the random and fixed effects are almost identical, indicating that the random effects are negligible for the prediction task.

**Fig. 7:**
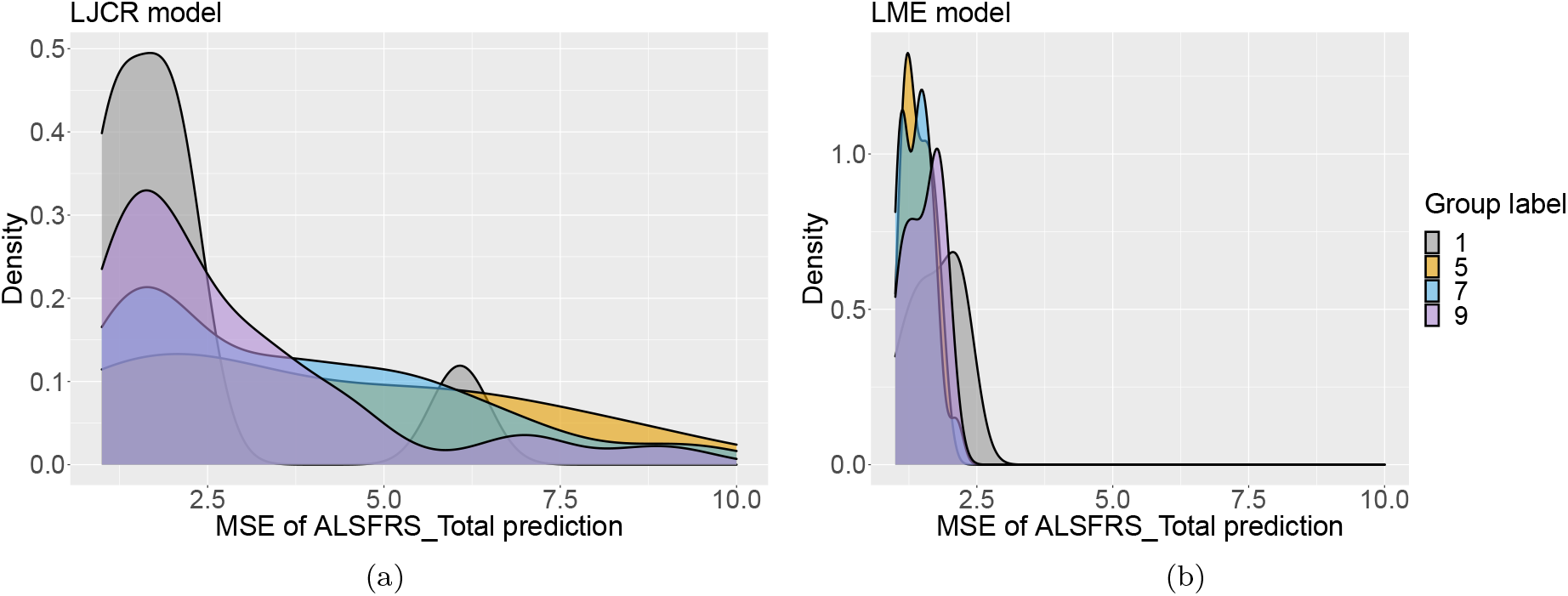
Distribution of ALSFRS prediction for each individual *i* and observation time *t* in different group labels ({1,5,7,9}).

**Fig. 8:**
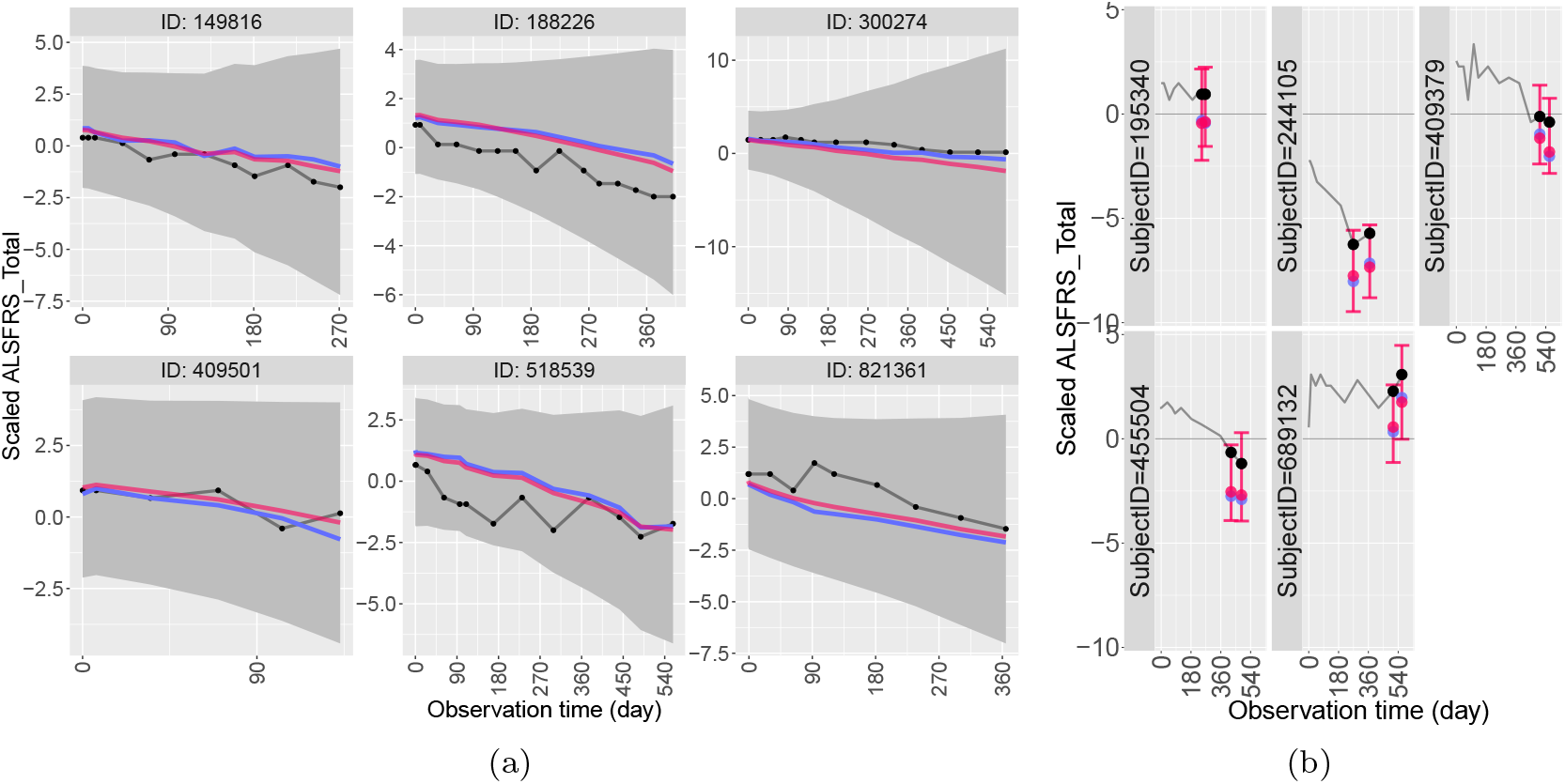
Performance of the LJCR model on ALS PRO-ACT data in predicting value of ALSFRS Total, when number of optimal groups is fixed and equal to *K*_optimal_ = 9. In each case, we compare performance of the LJCR model without taking the random effect parameters into account (in red), with the linear mixed effect model prediction (in blue). (a) Prediction for unseen individuals over a test set of individuals of size *n* = 50. The black line represents the true values of ALSFRS Total and the gray ribbon area shows the 95% confidence interval of the LJCR model when taking the random effect parameters into account. (b) Prediction for new datapoints of seen individuals over a test set of observation time-points of size 2. The test is executed on 5 randomly selected individuals. The black line represents the true values of ALSFRS Total and error bars show the 95% confidence interval of the LJCR model when taking the random effect parameters into account.

## 4. Discussion

The aim of this article was to introduce longitudinal joint cluster regression (LJCR) to detect latent group (cluster) structures within longitudinal data and predict personalised disease outcomes informed by these latent structures. Latent group structure plays a key role in modern data-intensive applications as it can strongly confound estimates and lead to practical difficulties if ignored.

Latent group structures are modelled using a class of Gaussian mixture models that couple together the multivariate distribution of the covariates and response. This is different from classical mixture regression approaches, which focus on the distribution of the dependent variable only. Our approach could be further extended to the non-parametric realm using e.g. a Dirichlet process formulation [Hannah et al., 2011, Liverani et al., 2015]. This would also remove the need for determining the optimal number of clusters. To avoid excessive computational costs, we have not pursued this approach here.

We model the longitudinal dynamics of each individual using a random effect intercept and slope model. The inference is done via a profile likelihood approach that can handle high-dimensional covariates by incorporating sparsity assumptions via ridge penalization. While l1 penalisation is possible in the mixed model paradigm [Schelldorfer et al., 2011], this comes with computational disadvantages, and the benefit of additional sparsity obtained by setting some parameters to zero is not clear; in previous work [Dondelinger et al., 2020], l2 penalisation led to improved predictions in some settings.

We have compared the performance of the LJCR model with an alternative method based on k-means [Genolini and Falissard, 2010] under a scenario where we vary the sample size and the number of covariates. It was shown that the LJCR outperforms this method, both in prediction error for the response variable (benefitting from modeling longitudinal dynamics via the random effect parameters), and prediction error for the fixed effect parameters in the high-dimensional case (benefitting from incorporating ridge penalization).

An alternative method is the one described in Bruckers et al. [2016], which uses a latent growth model for the longitudinal data. It is worth mentioning that Bruckers et al. [2016], like most conventional mixture model approaches, only relies on conditional distribution of responses *Y* |*X*, disregarding any signal arising from the distribution of feature matrix *X* itself. This is one of the key differences between the LJCR method and the other standard models as we also incorporate estimation of the distribution of *X* via a graphical lasso approach.

We applied LJCR to a cohort of patients with ALS disease to find the latent subtypes (groups) within the study. Our approach detected 9 group labels in total, with 4 groups hosting the largest population sizes. Note that we are not claiming this as a ground truth for the homogeneous groups within the dataset, but rather an estimate based on our linear mixed model approach for the dynamics within each mixture component. An interesting extension for our work would be to consider non-linear dynamics for the longitudinal model.

We evaluated the prediction performance on our real-world dataset for each of the larger groups, and found that post-inference refitting of a standard linear mixed model improves prediction error. As we do not have a gold standard for group membership, we investigate the group label assignments derived by the LJCR algorithm informally by looking at the group characteristics and interpreting the clinical and biochemical variables identified as important via the group-specific fixed effects. Further investigations should focus on confirmatory studies to establish whether these variables have a causal effect on disease progression in subsets of patients.

## Data Availability

The ALS PRO-ACT data is publicly available.
Parkinson's disease data (PPMI) can be found at the following link

https://nctu.partners.org/ProACT/

http://www.ppmi-info.org/access-data-specimens/download-data/

## References

Nazem Atassi, James Berry, Amy Shui, Neta Zach, Alexander Sherman, Ervin Sinani, Jason Walker, Igor Katsovskiy, David Schoenfeld, Merit Cudkowicz, et al. The pro-act database: design, initial analyses, and predictive features. Neurology, 83(19):1719–1725, 2014.

Liesbeth Bruckers, Geert Molenberghs, Pim Drinkenburg, and Helena Geys. A clustering algorithm for multivariate longitudinal data. Journal of Biopharmaceutical Statistics, 26(4): 725–741, 2016.

Oleg Butovsky, Shafiuddin Siddiqui, Galina Gabriely, Amanda J Lanser, Ben Dake, Gopal Murugaiyan, Camille E Doykan, Pauline M Wu, Reddy R Gali, Lakshmanan K Iyer, et al. Modulating inflammatory monocytes with a unique microrna gene signature ameliorates murine als. The Journal of clinical investigation, 122(9):3063–3087, 2012.

Patrick Danaher, Pei Wang, and Daniela M Witten. The joint graphical lasso for inverse covariance estimation across multiple classes. arXiv:1111.0324, November 2011.

David L Davies and Donald W Bouldin. A cluster separation measure. IEEE transactions on pattern analysis and machine intelligence, (2):224–227, 1979.

Frank Dondelinger, Sach Mukherjee, and Alzheimer’s Disease Neuroimaging Initiative. The joint lasso: high-dimensional regression for group structured data. Biostatistics, 21(2):219–235, 2020.

L Dupuis, P Corcia, A Fergani, J-L Gonzalez De Aguilar, D Bonnefont-Rousselot, R Bittar, D Seilhean, J-J Hauw, L Lacomblez, J-P Loeffler, et al. Dyslipidemia is a protective factor in amyotrophic lateral sclerosis. Neurology, 70(13):1004–1009, 2008.

Brian S Everitt, S Landau, and M Leese. Cluster analysis arnold. A member of the Hodder Headline Group, London, pages 429–438, 2001.

Jerome Friedman, Trevor Hastie, and Robert Tibshirani. Sparse inverse covariance estimation with the graphical lasso. Biostatistics, 9(3):432–441, 2008.

Jerome Friedman, Trevor Hastie, Rob Tibshirani, and Maintainer Rob Tibshirani. Package ‘glasso’, 2015.

Christophe Genolini and Bruno Falissard. Kml: k-means for longitudinal data. Computational Statistics, 25(2):317–328, 2010.

Lauren A Hannah, David M Blei, and Warren B Powell. Dirichlet process mixtures of generalized linear models. J. Mach. Learn. Res., 12(6), 2011.

Trevor Hastie. glmnet v4. 0-2. 2020.

Clifford M Hurvich and Chih-Ling Tsai. Regression and time series model selection in small samples. Biometrika, 76(2):297–307, 1989.

Salvatore Ingrassia, Simona C Minotti, and Giorgio Vittadini. Local statistical modeling via a cluster-weighted approach with elliptical distributions. Journal of classification, 29(3):363–401, 2012.

Kalpana D Joshi and PS Nalwade. Modified k-means for better initial cluster centres. International Journal of Computer Science and Mobile Computing, IJCSMC, 2(7):219–223, 2013.

Abbas Khalili and Jiahua Chen. Variable selection in finite mixture of regression models. Journal of the american Statistical association, 102(479):1025–1038, 2007.

N M Laird and J H Ware. Random-effects models for longitudinal data. Biometrics, 38(4): 963–974, December 1982.

Silvia Liverani, David I Hastie, Lamiae Azizi, Michail Papathomas, and Sylvia Richardson. PRe-MiuM: An R package for profile regression mixture models using dirichlet processes. J. Stat. Softw., 64(7):1–30, March 2015.

Jessica Mandrioli, Sara Biguzzi, Carlo Guidi, Elisabetta Sette, Emilio Terlizzi, Alessandro Ravasio, Mario Casmiro, Fabrizio Salvi, Rocco Liguori, Romana Rizzi, et al. Heterogeneity in alsfrs-r decline and survival: a population-based study in italy. Neurological Sciences, 36(12): 2243–2252, 2015.

G McLachlan. Peel., d.(2000). finite mixture models.

Hiroshi Mitsumoto, Diana C Garofalo, Regina M Santella, Eric J Sorenson, Björn Oskarsson, J americo M Fernandes Jr, Howard Andrews, Jonathan Hupf, Madison Gilmore, Daragh Heitzman, et al. Plasma creatinine and oxidative stress biomarkers in amyotrophic lateral sclerosis. Amyotrophic Lateral Sclerosis and Frontotemporal Degeneration, 21(3-4):263–272, 2020.

Konstantinos Perrakis, Frank Dondelinger, and Sach Mukherjee. Latent group structure and regularized regression. arXiv preprint arXiv:1908.07869, 2019.

José Pinheiro and Douglas Bates. Mixed-effects models in S and S-PLUS. Springer Science & Business Media, 2006.

José C Pinheiro and Douglas M Bates. Mixed-Effects Models in S and S-PLUS. Springer, New York, NY, 2000.

Siddheswar Ray and Rose H Turi. Determination of number of clusters in k-means clustering and application in colour image segmentation. In Proceedings of the 4th international conference on advances in pattern recognition and digital techniques, pages 137–143. Calcutta, India, 1999.

Joseph L Schafer. Some improved procedures for linear mixed models. Submitted to Journal of, 1998.

Jürg Schelldorfer, Peter Bühlmann, and Sara Van de Geer. Estimation for High-Dimensional linear Mixed-Effects models using l1-penalization. Scand. Stat. Theory Appl., 38(2):197–214, 2011.

Gideon Schwarz et al. Estimating the dimension of a model. The annals of statistics, 6(2): 461–464, 1978.

Nicolas Stådler, Peter Bühlmann, and Sara Van De Geer. l1-penalization for mixture regression models. Test, 19(2):209–256, 2010.

Harini Suresh, Jen J Gong, and John V Guttag. Learning tasks for multitask learning: Heterogenous patient populations in the ICU. In Proceedings of the 24th ACM SIGKDD International Conference on Knowledge Discovery & Data Mining, KDD’18, pages 802–810, New York, NY, USA, July 2018. Association for Computing Machinery.

Bart Swinnen and Wim Robberecht. The phenotypic variability of amyotrophic lateral sclerosis. Nature Reviews Neurology, 10(11):661, 2014.

Harry Joe Weiliang Qiu. Package ‘clustergeneration’, 2015.

Jianpeng Xu, Jiayu Zhou, and Pang-Ning Tan. FORMULA: FactORized MUlti-task LeArning for task discovery in personalized medical models. In Proceedings of the 2015 SIAM International Conference on Data Mining, Proceedings, pages 496–504. Society for Industrial and Applied Mathematics, June 2015.

Jing Zhao. Package ‘linear mixed models (lmm) package v1.3’, 2020.

